# Long lockdowns and rainy days: Modeling the interactive roles of weather, behavior, and restrictions in COVID-19 transmission in the Netherlands

**DOI:** 10.1101/2021.03.16.21253684

**Authors:** Ondrej Mitas, Alinda Kokkinou

## Abstract

Extant research on the role of weather in COVID-19 has produced ambiguous results and much methodological debate. Following advice emerging from this methodological debate, we take a step further in modeling effects of weather on COVID-19 spread by including interactions between weather, behavior, baseline cases, and restrictions in our model. Our model was based on secondary infection, hospitalization, restriction, weather, and mobility data per day nested with safety region in the Netherlands. Our findings show significant but inconsistent interactions. The robust effects of weather on COVID-19 spread persisted over and above these interactions, highlighting the need to account for weather with nuance and caution in public policy, communication, and forecasting.

## Introduction

There is much debate in the scientific (Lasisi & Eluwole, 2021; Pedrosa, 2020; Sahoo, Powell, Mittal, & Garg, 2020) and popular (Hart, 2021) literature about the effects of weather on COVID-19 transmission. Many European countries have experienced clear first and second waves of peaking COVID-19 infections coinciding with relatively cold and wet spring and autumn weather, respectively, while other regions have experienced vigorous outbreaks even during very warm conditions. Social distancing and lockdown measures rarely take weather and seasonal differences into account. Furthermore, while leaders’ communication and example-setting related to the pandemic clearly affects behavior, the effects of weather are generally absent from these messages as well. Medical experts generally send the message that effects of weather on COVID-19 spread are not well known (Hart, 2021) and that extant research varies dramatically in quality, especially in terms of interactions with extraneous variables (Zaitchik et al., 2020). While numerous scientific models linking weather to COVID-19 growth rates exist, they often comprise mere bivariate analyses between weather and growth rates “leading to potentially spurious conclusions “(Carlson, Gomez, Bansal, & Ryan, 2020), or at best multivariate models in which other variables are controlled for (Pedrosa, 2020).

In the present analysis, we analyze spread of COVID-19 infections in the Netherlands within the 25 Dutch safety regions using a multilevel longitudinal model, where we not only control for temporal influences, but also model interactions between weather, restrictions, and recorded behavior. This level of modeling complexity is needed to advise the public in a way that accounts for the effects of weather, in line with the calls by Sahoo et al. (2020) and (Zaitchik et al., 2020) to study the effects of weather on the pandemic in more depth with interaction and time-lag effects transparently accounted for.

Our theoretically driven model assumes that newly reported cases on any given day are driven primarily by the extant cases from which new infections are obtained, taking a 6-day incubation period. Rather than modeling a growth rate as the outcome, which ignores the psychological effect of case numbers and pandemic spread communicated in the media, we used the cases on a given day, as well as the contextual variables we were interested in––weather, restrictions, and behavior––as predictors of new cases six days later. We similarly modeled hospitalizations on a 10-day lag, which allowed us to include data obtained before testing became widely available in the Netherlands on 1 June 2020. Our modeling approach assumed that weekend days and holidays needed to be controlled for, and the number of cases on day 0 should be allowd to interact with weather and restrictions. We also posited that restrictions and weather affect not only case numbers but growth, implying 3-way interactions between weather, restrictions, and day 0 cases.

The discussion about the role of weather in spreading the virus partly focuses on the distinction between the fitness and transmissibility of the virus at colder temperatures (Carlson et al., 2020), and the change in human behavior when the weather is not pleasant enough to gather outside. To shed light on this ambiguity, we initially ran models without any measures of human behavior. These were later added as potential mediators. If the effects of weather-restriction-case interactions were attenuated by the addition of behavioral predictors, it would suggest that COVID-19 spreads more readily in unpleasant weather largely by driving people to gather indoors. Conversely, the absence of such a mediation effect would point to the various hypotheses suggesting that colder, wetter weather facilitates fitness and transmissibility of the virus itself.

It is important to note that this research note is not intended to give a full review of previous research on the topic, which is available elsewhere (Sahoo et al., 2020), to fully discuss the theoretical implications of our findings, nor to provide forecasts which can be used to make policy based on meteorological inputs. Rather, we want to report concisely on a new model which adds depth to previous methodological perspectives to the weather-COVID-19 link. We aim not to definitively quantify this connection, but to inspire researchers to continue this effort and to take multivariate interactive perspectives on epidemiological effects of weather seriously in knowledge- and decision-making processes. As such, we follow the advice emerging from the symposium on Climatological, Meteorological, and Environmental factors in the COVID-19 pandemic to:

> account for relevant non-environmental predictors; to justify the data quality and relevance of the selected response variable; to be clear about the epidemic phase being tested (including time lags); to spatially and temporally align epidemiological and climate, meteorological and environmental data; and to distinguish between analyses that are suited to describe observed relationships and those that have been confirmed to provide skillful prediction and forecast (Zaitchik et al., 2020).

The latter point is particularly significant, wherein our analysis addresses observed relationships from previous data, and is *not* a forecasting model.

## Methods

### Data

The present analysis is built on secondary data. Data on daily COVID-19 cases and hospitalizations was obtained from the Dutch RIVM (*Rijsinstituut voor Volksgezondheid en Milieu*, National Institute for Public Health and Environment), which is entrusted by the government to track the pandemic in the Netherlands (RIVM, 2021). These data are reported by safety region, a geographical classification used in the Netherlands as part of crisis management. Behavior was operationalized by mobility. The Google Community Mobility Reports were used (Google LLC, 2021). These datasets, compiled using anonymized data, show how visits and length of stay changed during the pandemic as compared to a baseline. The data are aggregated according to public transport, work, retail, and residential categories. Other categories such as parks and natural areas were excluded from analysis due to large amounts of missing data. The Google Community Mobility Reports used a different geographical classification to aggregate data than the safety regions used by the Dutch RIVM. There was overlap between six safety regions and mobility regions. However, for the remaining safety regions, typically more densely populated, there was no overlap, and the most prominent cities were used as a proxy. Weather data were accessed from the Dutch KNMI (2021, Royal Netherlands Meteorological Institute). For each safety region, a single weather station was selected. However, for two pairs of safety regions this was not possible. Therefore VR09 (Utrecht) and VR14 (Gooi en Vechtstreek) share weather data from station 260 (De Bilt) and VR08 (Gelderland-Zuid) and VR18 (Zuid-Holland-Zuid) share weather data from station 356 (Herwijnen). Data on dates when measures became effective, or lifted, were obtained from the official government organization (Rijksoverheid, 2021) and reports of Dutch public broadcasting (Nationale Omroep Stichting).

|  | **Safety Region** | **Weather Station** | **Mobility Region** |
| --- | --- | --- | --- |
| VR01 | Groningen | 280 Eelde | NL-GR |
| VR02 | Friesland | 270 Leeuwarden | NL-FY |
| VR03 | Drenthe | 279 Hoogeveen | NL-DR |
| VR04 | IJsselland | 278 Heino | Zwolle |
| VR05 | Twente | 290 Twenthe | Enschede |
| VR06 | Noord- en Oost-Gelderland | 283 Hupsel | Apeldoorn |
| VR07 | Gelderland-Midden | 275 Deelen | Arhem |
| VR08 | Gelderland-Zuid | 356 Herwijnen | Nijmegen |
| VR09 | Utrecht | 260 De Bilt | NL-UT |
| VR10 | Noord-Holland-Noord | 235 De Kooy | Alkmaar |
| VR11 | Zaanstreek-Waterland | 249 Berkout | Zaanstad |
| VR12 | Kennemerland | 275 Wijk aan zee | Haarlem |
| VR13 | Amsterdam-Amstelland | 240 Schiphol | Government of AMS |
| VR14 | Gooi en Vechtstreek | 260 De Bilt | Hilversum |
| VR15 | Haaglanden | 330 Hoek van Holland | The Hague |
| VR16 | Hollands-Midden | 348 Cabauw | Leiden |
| VR17 | Rotterdam-Rijnmond | 344 Rotterdam | Government of Rotterdam |
| VR18 | Zuid-Holland-Zuid | 356 Herwijnen | Dordrecht |
| VR19 | Zeeland | 310 Vlissingen | NL-ZE |
| VR20 | Midden- en West-Brabant | 350 Gilze | Breda |
| VR21 | Brabant-Noord | 375 Volkel | s-Hertogenbosch |
| VR22 | Brabant-ZuidOost | 370 Eindhoven | Eindhoven |
| VR23 | Limburg-Noord | 391 Arcen | Venlo |
| VR24 | Limburg-Zuid | 380 Maastricht | Maastricht |
| VR25 | Flevoland | 269 Lelystad | NL-FL |

### Analysis

Because our data record a longitudinal change process nested within regional units, a multilevel random intercept model was deemed appropriate. Such a model is based on a linear modeling logic but allows the intercept to vary randomly between groups in the data, in this case the 25 safety regions of the Netherlands. The outcome variables in the models were log-transformed daily new cases recorded 6 days after the date of the predictor variables, and log-transformed daily new hospitalizations recorded 10 days after the date of the predictor variables, respectively. We built our models by entering day of weekend (Friday, Saturday, and Sunday) and holiday dummy variables as controls, as well as 3-way interactions between a weather index, log-transformed daily new cases or hospitalizations, and dummy-coded restrictions, including closing restaurant and bars, closing non-essential shops, mandatory indoor facemasking, and a curfew. The weather index (Cronbach alpha > 0.71; Revelle’s Omega Total = 0.93) comprised an average of z-standardized high temperature, low temperature, hours of sunshine, hours of precipitation (reverse coded), and millimeters of precipitation (reverse coded). A subsequent model included four-way interactions including a mobility index (Cronbach alpha > 0.82; Revelle’s Omega Total > 0.93) with an average of retail, work, transit, and residential (reverse coded) mobility. These four-way interactions were not significant at the 0.05 alpha level, and were thus trimmed out of the model.

Finally, instead of interacting mobility with other variables, we included the mobility index as well as each mobility measure separately as a mediating covariate in two separate versions of our models. Due to having lower Bayesian Information Criteria, which penalize excessive predictors, we chose the models with separate mobility measures over those with a single mobility index. The coefficients of weather-restriction-case 3-way interactions did not decrease as a result of including mobility predictors. Some of the mobility predictors were significant on their own, however, and were thus retained in our definitive models (Tables 1 and 2).

**Table 1.** Model of daily new cases 1 June 2020 - 2 Feb 2021

| Predictor | Estimate | Std. Error |  |
| --- | --- | --- | --- |
| **(Intercept)** | **0.1774781** | **0.0394509** | **\*\*\*** |
| Friday | -0.0187417 | 0.0223123 |  |
| **Saturday** | **0.0917894** | **0.0311371** | **\*\*** |
| Sunday | 0.0517782 | 0.0327639 |  |
| Holidays | -0.0693239 | 0.0475004 |  |
| Weather | 0.0491801 | 0.0293102 |  |
| **Closing bars/restaurants** | **0.7013477** | **0.0946091** | **\*\*\*** |
| **ln(Total Cases)** | **0.9624250** | **0.0067444** | **\*\*\*** |
| Curfew | 0.1748585 | 0.9221829 |  |
| Closing shops | -0.1554003 | 0.6673706 |  |
| Face mask requirement | 0.8385140 | 0.5911271 |  |
| **Residential mobility** | **-0.1667361** | **0.0207708** | **\*\*\*** |
| **Work mobility** | **-0.1814930** | **0.0180943** | **\*\*\*** |
| Retail mobility | -0.0005475 | 0.0182446 |  |
| **Transit mobility** | **0.1108813** | **0.0156779** | **\*\*\*** |
| **Weather x Closing bars/restaurants** | **-0.8649414** | **0.1706616** | **\*\*\*** |
| **Weather x ln(Total Cases)** | **-0.0846045** | **0.0095868** | **\*\*\*** |
| **Closing bars/restaurants x ln(Total Cases)** | **-0.1318889** | **0.0177091** | **\*\*\*** |
| Weather x curfew | 0.6511103 | 1.0348254 |  |
| ln(Total Cases) x curfew | -0.0749759 | 0.1798114 |  |
| Weather x Closing most shops | 0.1372678 | 0.8963203 |  |
| ln(Total Cases) x Closing most shops | -0.0223474 | 0.1197400 |  |
| Weather x Face mask requirement | 0.4718478 | 0.8294427 |  |
| ln(Total Cases) x Face mask requirement | -0.0758725 | 0.1073570 |  |
| **Weather x Closing bars/restaurants x ln(Total Cases)** | **0.1994290** | **0.0342122** | **\*\*\*** |
| Weather x ln(Total Cases) x curfew | -0.1360893 | 0.2008918 |  |
| Weather x ln(Total Cases) x Closing most shops | -0.0127719 | 0.1614483 |  |
| Weather x ln(Total Cases) x Face mask requirement | -0.0576164 | 0.1509528 |  |

Note: P-values according to Satterthwaite t-values coded as follows.

**Table 2.**
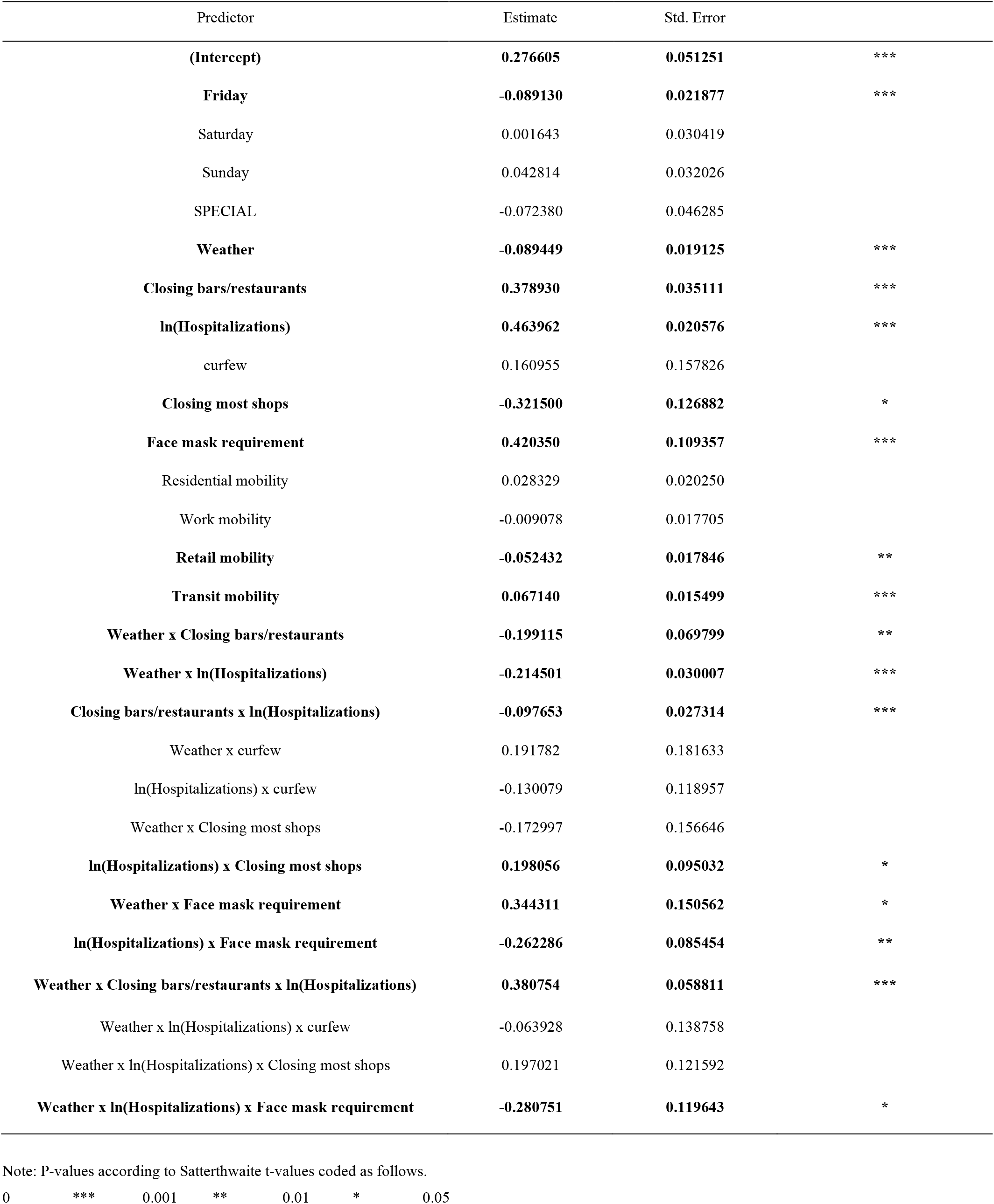
Model of daily new hospitalizations 27 Feb 2020 - 2 Feb 2021

## Results

Due to the multi-parameter nature of 3-way interactions, the findings can only be interpreted graphically, not based on signs and magnitudes of individual parameters. For the model of COVID-19 cases based on 1 June 2020 - 1 February 2021 data, only the restaurant and bar-closure x weather x cases 3-way interaction was significant. Figure 1 illustrates this interaction. We assumed a mean level of daily new day 0 cases and modeled the change from this baseline compared to daily new cases 6 days later for mean, mean-1SD (“unpleasant”), and mean+1SD (“pleasant”) levels of our weather index at 0 (bars and restaurants open) and 1 (bars and restaurants closed) levels of restriction. The graph shows a clear downward trend in case growth from unpleasant to pleasant weather, and shows that this pattern is slightly steeper, thus the effect of weather accentuated, when bars and restaurants are closed.

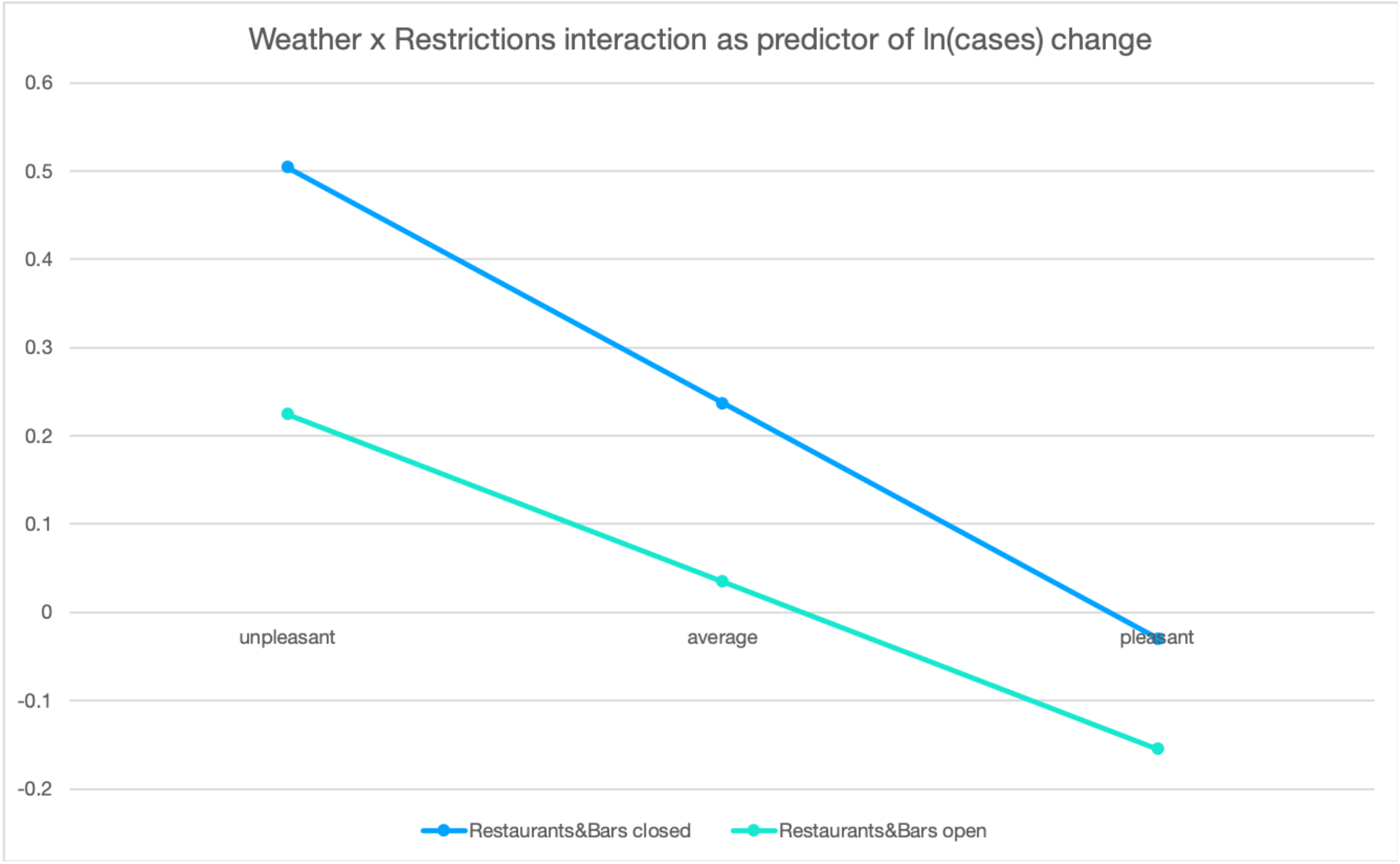

The same 3-way interaction was significant in the model of COVID-19 hospitalizations between 1 April 2020 and 1 February 2021. We also found a second significant 3-way interaction, this time concerning the restriction of facemasks required in indoor public spaces (including restaurants as well as shops in the Netherlands). Because facemasks were only required while restaurants were closed, we graphed 3 instances of restrictions at unpleasant, pleasant, and average levels of weather: bars and restaurants open, bars and restaurants closed but facemasking not required, and both restrictions in place. The pattern shows similarly steep slopes in hospitalization growth from unpleasant to pleasant weather for the least and most restricted situation. The effect of weather on hospitalization growth was less strong when restaurants were closed, but facemasking was not required. This concerned the period between April and June 2020, and a second period of six weekend between October and November 2020.

In sum, there is an ambiguous interaction between closing restaurants and weather as contributors to growth in spread of COVID-19. Nevertheless, there was no interaction with restriction or behavior that attenuated the effect of weather to the point where its facilitating effects could be neutralized. The effects observed were not reduced by the inclusion of mobility predictors, suggesting that unpleasant weather supports the fitness and transmissibility of the virus above and beyond the effect that weather has on individuals’ behavior.

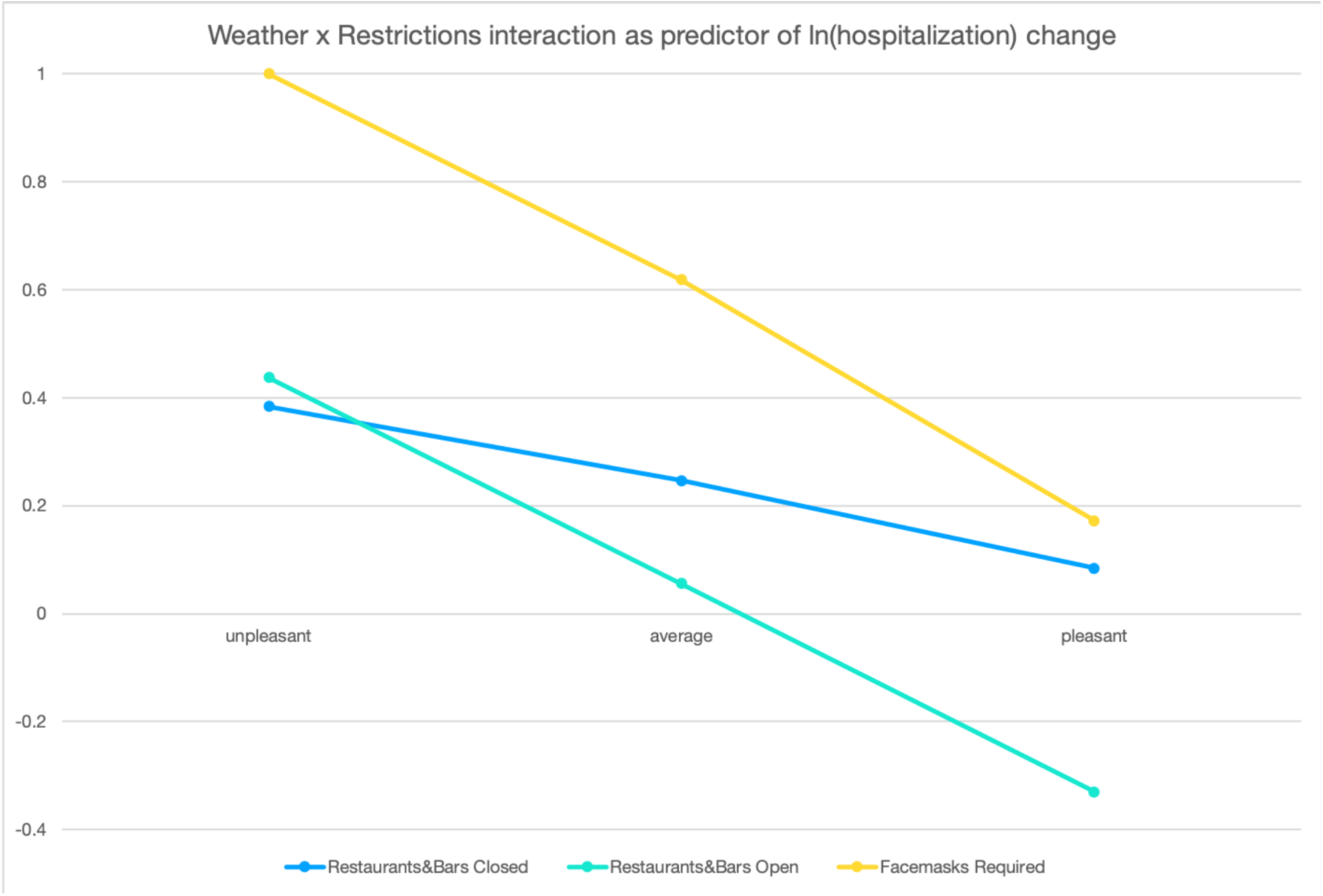

Considering that, as expected, weather had a substantial influence on increase in COVID-19 cases and hospitalizations, we also used the statistically significant parameters in our model to quantify the difference between relatively pleasant (mean + 1 standard deviation) and unpleasant (mean - 1 standard deviation) days. During the time period in the cases dataset (1 June 2020 - 2 February 2021), a pleasant day would feature a high temperature of 23.9 C, a low temperature of 13.7 C, 9 hours of sun, and no precipitation. An unpleasant day would feature a high temperature of 8.3 C, a low temperature of 2.5 C, less than an hour of sun, and 7.4 mm of precipitation over 5.1 hours. For a mean level of daily new cases (3742), our model estimates that difference between a pleasant- and unpleasant-weather day corresponded to a difference of 313 cases six days later per day nationally. With the effect of restrictions removed from the model, this difference was higher, namely 365 cases per day nationally.

During the time period in the hospitalizations dataset (1 March 2020 - 2 February 2021), a pleasant day would feature a high temperature of 23.1 C a low of 12.7 C, 10.2 hours of sun, and no precipitation. An unpleasant day would feature a high temperature of 12.7 C a low of 1.7 C, 1 hour of sun, and 6.5mm of precipitation over 4.8 hours. For a mean level of daily new hospitalizations nationally (66), the difference between a pleasant- and unpleasant-weather day corresponded with a difference of 1 hospitalization per day nationally 10 days later. With the effect of restrictions removed, this difference rose to 12 hospitalizations per day.

## Conclusion

The COVID-19 pandemic has caused untold grief and damage to health as well as society. Well-meaning governments have undertaken a variety of restrictions to control the spread of the virus, with varying success depending on compliance, fear, and other social factors. It is noted that some restrictions have snowballed into new health crises of their own, such as mental health care for young people in the Netherlands (Daniels, 2021). Public policy instruments to combat the pandemic are only as effective as the knowledge on which they are based. The knowledge about how weather affected the spread of COVID-19––especially in interaction with restrictions and behavior––has been lacking, ambiguous, or contradictory, depending on the source (Zaitchik et al., 2020). We found that weather had a substantial effect on the spread of COVID-19 in a way that previous restrictions in the Netherlands failed to neutralize. Whether “summer weather is unlikely to be protective” (Carlson et al., 2020) is disputable. Our findings do not suggest that pleasant weather alone prevents infection of a specific individual, but on an epidemiological level, it is protective in the sense of somewhat reducing virus transmission on a population level. This reduction occurred in concert with public restrictions and communication, however. No matter how pleasant, weather does not render social distancing, masking, or some extent of lockdowns unnecessary. A more useful conclusion from the findings is that unpleasant weather facilitated the spread of the virus over and above day of weekend, holidays, restrictions, and mobility. In sum, pleasant weather does not obviate the need for restrictions, but restrictions alone are insufficient to negate the epidemiological effect of unpleasant weather. Our results are consistent with the account that unpleasant weather facilitates COVID-19 spread by increasing fitness and transmissibility of the virus over and above the effects of weather on human behavior (e.g., gathering indoors on rainy days), though not conclusive on this point.

Thus, we urge leaders and policy-makers to take seasonal patterns and weather forecasts into account when selecting restriction, communication, and leadership strategies. Furthermore, seasonal patterns can contribute some much-needed predictability to these strategies, which may contribute to compliance. Finally, the reduction in cases that may occur as summer weather approaches and the weather improves should not lead to complacency. Instead, policymakers should use this respite to take necessary measures, such as vaccinations, to ensure that return to inclement autumn weather does not, once again, facilitate conditions for accelerated COVID-19 spread.

### Limitations

Carlson et al. (2020) urge researchers to “identify the effects of weather on processes like disease transmission accounting for confounders, lags, and bias in climate and disease data” (p. 11). With our model, we have attempted to heed this advice, but have only scratched the surface in terms of potential model complexity and temporal dynamics. This analysis was meant to offer a deeper epidemiological look at the effects of weather on the spread of COVID-19, not to definitively quantify or forecast how much changes in weather lead to changes in the spread of the virus. As such, our approach comes with certain limitations. Our model is theory-driven.

Thus, we did not model highly complex patterns of interaction which could, for example, be uncovered by machine learning approaches. Furthermore, we worked with available data. These do not include self-report questionnaires or other measures of underlying psychological processes. Such psychological processes could explain why a person chooses to undertake risky behaviors within (or in violation of) restrictions, and may themselves have more complex interactions with the weather. For example, our predictor of baseline-level (day 0) cases models both sources of virus transmission and the psychological effect of new case counts communicated in the media. To capture both of these effects using the baseline case count is a rather blunt instrument, however. An improvement would be to use self-response questionnaires to parse the psychological mechanisms at play, while using the reproduction factor or another growth metric as the outcome variable of the model.

### Suggestions for future research

Considering the limited scope of the present data and analysis, there are relatively accessible approaches that, in future research, could greatly increase the validity of models we have presented. Concretely, we make two suggestions. First, we used the same time lag for conditions in which COVID-19 transmission takes place, and for the baseline level of infections on which that transmission builds. In our analysis of reported cases, for example, we thus assume that people transmit the virus to others and receive positive test results the same day. Future research should challenge this assumption and, if the data supports it, adopt different time lags for baseline case or hospitalization numbers and environmental as well as behavioral variables. Some studies have also used 3-day averages as a smoothing function, an approach worth considering.

Second, our model was an observational explanatory model and is not intended for forecasting. The importance of weather we uncovered, however, points to the relevance of including weather in models which *are* intended to quantitatively forecast future cases.

Improving the accuracy of forecasting models is a priority to target restrictions and public communication as precisely as possible. Critically examining weather variables and their interactions with other predictors can play a crucial role in improving COVID-19 forecasting.

## Supporting information

Hospitalization Data

Case Data

Example analysis script

## Data Availability

The data file will be uploaded alongside the MS.

